# Home stay reflects symptoms severity in major depressive disorder: A multicenter observational study using geolocation data from smartphones

**DOI:** 10.1101/2021.02.10.21251512

**Authors:** Petroula Laiou, Dzmitry A. Kaliukhovich, Amos Folarin, Yatharth Ranjan, Zulqarnain Rashid, Pauline Conde, Callum Stewart, Shaoxiong Sun, Yuezhou Zhang, Faith Matcham, Alina Ivan, Grace Lavelle, Sara Siddi, Femke Lamers, Brenda W.J.H. Penninx, Josep Maria Haro, Peter Annas, Nicholas Cummins, Srinivasan Vairavan, Nikolay V. Manyakov, Vaibhav A. Narayan, Richard Dobson, Matthew Hotopf, on behalf of the RADAR-CNS consortium

**Affiliations:** Department of Biostatistics and Health Informatics, Institute of Psychiatry, Psychology and Neuroscience, King’s College London, London, UK; Janssen Pharmaceutica NV, Beerse, Belgium; Institute of Health Informatics, University College London, London, UK; South London and Maudsley NHS Foundation Trust, London, UK; Psychological Medicine, Institute of Psychiatry, Psychology and Neuroscience, King’s College London, London, UK; Parc Sanitari Sant Joan de Déu, Fundació Sant Joan de Déu, Centro de Investigación Biomédica en Red Salud Mental, Universitat de Barcelona, Barcelona, Spain; Department of Psychiatry and Amsterdam Public Health Research Institute, Amsterdam UMC, Vrije Universiteit, Amsterdam, the Netherlands; H. Lundbeck A/S, Valby, Denmark; Chair of Embedded Intelligence for Health Care and Well-being, University of Augsburg, Augsburg, Germany; Janssen Research & Development, LLC, Titusville, NJ, USA

**Keywords:** major depressive disorder, PHQ-8, smartphone, GPS, home stay

## Abstract

Most smartphones and wearables are nowadays equipped with location sensing (using Global Positioning System and mobile network information) that enable continuous location tracking of their users. Several studies have reported that the amount of time an individual experiencing symptoms of Major Depressive Disorder (MDD) spends at home a day (i.e., home stay), as well as various mobility related metrics, are associated with symptom severity in MDD. Due to the use of small and homogeneous cohorts of participants, it is uncertain whether the findings reported in those studies generalize to a broader population of individuals with the MDD symptoms. In the present study, we examined the relationship between overall severity of the depressive symptoms, as assessed by the eight-item Patient Health Questionnaire (PHQ-8), and median daily home stay over the two weeks preceding the completion of a questionnaire in individuals with MDD. We used questionnaire and geolocation data of 164 participants collected in the observational Remote Assessment of Disease and Relapse – Major Depressive Disorder (RADAR-MDD) study. Participant age and severity of the MDD symptoms were found to be significantly related to home stay, with older and more severely affected individuals spending more time at home. The association between home stay and symptom severity appeared to be stronger on weekdays than on weekends. Furthermore, we found a significant modulation of home stay by occupational status, with employment reducing home stay. Our findings suggest that home stay is associated with symptom severity in MDD and demonstrate the importance of accounting for confounding factors in future MDD studies.

## Introduction

The World Health Organization ranks depression as the single largest contributor to global disability (World Health Organization 2017). People with major depressive disorder (MDD) often experience physical comorbidity (Cimpean and Drake 2011), loss of occupational function (Lerner et al. 2004) and low quality of life (Lenox-Smith et al. 2013). Furthermore, MDD is strongly associated with suicidal deaths and premature mortality (Cuijpers and Schoevers 2004). The process for MDD diagnosis and evaluation of symptoms severity is highly dependent on the subjective information that an individual under screening provides to a clinician, and it might suffer from recall bias.

The recent advances in digital technologies including smartphones and wearable devices enable the collection of a variety of data streams that can be utilized to objectively characterize an individual’s daily activity and physical condition (Callan et al. 2017). These data can be collected continuously, remotely and unobtrusively without affecting an individual’s daily routine and behavior. Importantly, analysis of such data could result in the development of new objective, quantifiable, cost effective, and viable digital markers of an individual’s behavioural, cognitive, and emotional state (Grist et al. 2017; Mohr et al. 2017; Rohani et al. 2018).

Several recent studies have demonstrated associations between the MDD symptoms and mobility patterns derived from mobile devices. For example, individuals with greater severity of the MDD symptoms were reported to make fewer transitions between locations of interest (i.e., those frequently visited in the past) and spend more time at home (Canzian and Musolesi 2015; Chow et al. 2017; Saeb et al. 2015, 2016; Wahle et al. 2016). Home stay, an indicator of social disengagement (Chow et. al 2017), was also reported to be significantly related to the severity of the MDD symptoms (Chow et al. 2017; Saeb et al. 2015, 2016).

Most studies used small and homogeneous cohorts of participants (e.g., university students) and were conducted over a short period (e.g., several weeks). In the present study, we examined the association between overall severity of the MDD symptoms and a measure of daily mobility patterns using data from a larger and more diverse group of participants collected in the Remote Assessment of Disease and Relapse – Major Depressive Disorder study (RADAR-MDD; Matcham et al. 2019). The RADAR-MDD study is an observational, longitudinal, prospective study that is currently being conducted at multiple clinical sites spread across several European countries and part of a wider program of research (RADAR-CNS; Remote Assessment of Disease and Relapse – Central Nervous System; http://radar-cns.org/) to explore the potential of wearable devices to help prevent and treat depression, multiple sclerosis and epilepsy. We used the eight-item Patient Health Questionnaire (PHQ-8; Kroenke et al. 2009) total score to measure severity of the MDD symptoms and home stay to describe an individual’s daily mobility patterns. Home stay was selected as an interpretable measure of mobility with previous evidence suggesting that it is related to the severity of the MDD symptoms (e.g., Chow et al. 2017; Saeb et al. 2015, 2016). Additionally, we examined whether the strength of the relationship changes from weekdays to weekends (i.e., modulated by changes in daily routine) and can be affected by an individual’ s demographics and quality of the acquired Global Positioning System (GPS) recordings. We hypothesised that higher levels of MDD as quantified by the PHQ-8 questionnaire would correspond to a prolonged home stay. In addition, we anticipated that a relationship between severity of MDD symptoms and home stay would be modulated by changes in daily routine from weekdays to weekends (Saeb et al. 2016).

## Materials and Methods

### Study population

Analysed participants were recruited to the RADAR-MDD study from November 2017 to November 2019. In addition, recruited participants were aged ≥ 18 years and had experienced at least two episodes of MDD in their lifetime, with the most recent within the last two years. The exclusion criteria included: lifetime history of bipolar disorder, schizophrenia, MDD with psychotic features, schizoaffective disorders; history of moderate to severe drug or alcohol dependence within six months prior to enrollment; history of a major medical disease which could impact upon the patient’s ability to participate in normal daily activities for more than two weeks; dementia; pregnancy. No limitations were applied regarding any treatment that the participants were receiving over the course of the study. Written consent was obtained previous to the enrollment session followed by collection of sociodemographic, social environment, medical history and technology use questionnaires, and the Lifetime Depression Assessment Self-Report (LIDAS; Bot et al. 2017). The participants with MDD were recruited at three clinical sites: King’s College London (KCL) in the UK, Vrije Universiteit Medisch Centrum (VUMC) in Amsterdam, the Netherlands, and Centro de Investigación Biomédica en Red (CIBER) in Barcelona, Spain (Table 1; Supplementary Table 1). More details on the study protocol can be found in Matcham et al. (2019).

**Table 1.** Dataset characteristics.

| Clinical site | KCL | CIBER | VUMC | All sites |
| --- | --- | --- | --- | --- |
| Characteristic |  |  |  |  |
| # Participants with both PHQ-8 and GPS data collected | 232 (57.9%) | 116 (28.9%) | 53 (13.2%) | 401 (100.0%) |
| # Participants with biweekly segments fulfilling the selection criteria | 109 (66.4%) | 38 (23.2%) | 17 (10.4%) | 164 (100.0%) |
| Of those, # female participants (% in a group) | 83 (76.1%) | 26 (68.4%) | 14 (82.4%) | 123 (75.0%) |
| Age (MED, SD, range), years | 46 (15.0)<br>18-73 | 54 (9.8)<br>27-71 | 33 (14.9)<br>19-69 | 48 (14.7)<br>18-73 |
| # Biweekly segments analyzed | 483 (62.8%) | 222 (28.9%) | 64 (8.3%) | 769 (100.0%) |
| Of those, for the employed participants | 277 (72.5%) | 64 (16.8%) | 41 (10.7%) | 382 (100.0%) |
| Of those, for the unemployed participants | 204 (53.0%) | 158 (41.0%) | 23 (6.0%) | 385 (100.0%) |

The number of participants and biweekly segments collected at each site was normalized by the corresponding total obtained by pooling data across the three sites (column “All sites”), with the resulting percentages being indicated in parentheses. Note that two biweekly segments from KCL had no data on occupational status. Abbreviations: # – number of, MED – median, SD – standard deviation.

### Data collection

We used the RADAR-base platform for data collection and storage (http://radar-base.org/; Ranjan et al. 2019). Participants with MDD were required to install several apps on their smartphones. The participants without a smartphone or with a non-Android device were provided with an Android smartphone and were required to use it throughout the study (Matcham et al. 2019). Remote monitoring technology (RMT) apps were used to collect data on participant severity of experienced MDD symptoms, self-esteem, cognitive functioning, voice audio sampling and brief in-the-moment assessments of daily life experiences. Specifically, every two weeks the participants were requested to fill in the PHQ-8 questionnaire in the RADAR-base active RMT app. The request notifications were sent out at a calendared time and remained active only on that day initially. The completion window was increased to three days to improve completion rates in April 2019.

The passive RMT apps ran in the background and required minimal input from participants. They collected data on participant physical (e.g., transitions in space) and socially relevant activity (e.g., number and duration of calls) as well as on some ambient factors (e.g., ambient noise and light). GPS location data were obfuscated by adding a fixed random number to the latitude and longitude of all GPS data points generated by a single participant (Figure 1). Accuracy of each acquired GPS data point, as provided by either a mobile network operator or GPS satellites, corresponded to one standard deviation of a bivariate normal distribution with equal variances along the two spatial dimensions centered at that point. GPS data points with the accuracy of larger than 20 meters were discarded from analyses (Supplementary Table 2). Sampling period of the GPS signal was set to either 5 (Depp et al. 2019; Saeb et al. 2015, 2016) or 10 (Barnett et al. 2018; Farhan et al. 2016a, 2016b) minutes throughout the study. However, the effective sampling period varied over time due to occasional signal loss and battery drain (i.e., signal undersampling). Other factors affecting the sampling period included occasional concurrent and asynchronous acquisition of geolocation data from both a mobile network operator and GPS satellites (i.e., signal oversampling).

**Figure 1.**
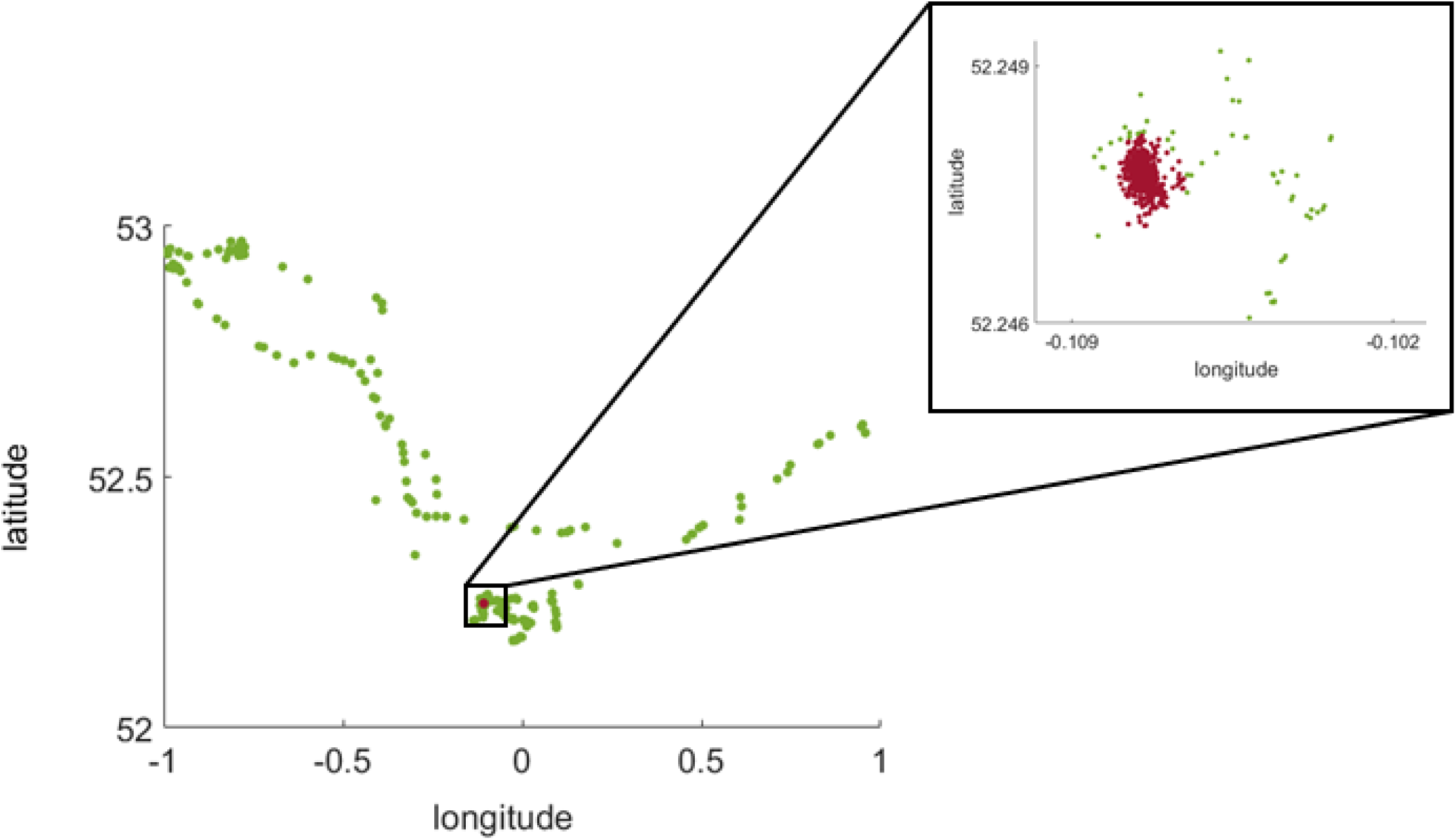
Exemplar geolocation data of a single study participant. GPS data were pooled over an exemplar biweekly segment. A single dot indicates a single acquired GPS location data point. The red dots denote individual’s home location, whereas the remaining dots are colored in green.

A single completed PHQ-8 questionnaire combined with the GPS data acquired over the two preceding weeks and obtained from the same participant is herein referred to as a biweekly segment. To ensure a high quality of the analyzed geolocation data, only biweekly segments that met the following criteria were analyzed: 14 days of GPS recordings available, and a daily median sampling period of the GPS signal ≤ 11 minutes, and the daily number of acquired GPS data points ≥ 48. The cut-off values were selected to maximize the volume of data available for analysis while preserving high quality of these data (Supplementary Tables 2, 3 and 4). Participants who declared their occupational status as a volunteer, student, caregiver, full- or part-time employee, or self-employed person were considered to be “employed”, while all other participants were considered to be “unemployed”. To avoid interference of the effect of the COVID-19 pandemic, only data collected before 1 January 2020 were analyzed.

For each day in a biweekly segment we computed the number of GPS data points collected for each of the 24 hours separately and over the entire day. Ideally, a GPS signal sampled uniformly at a period of 5 minutes would give 12 GPS data points an hour and a total of 288 GPS data points a day. We specified completeness of the daily data as a ratio between the actual number of GPS data points collected over a day and the expected one, as determined by a sampling period, with the values of 0 and 1 corresponding to an empty and a complete day of GPS recordings, respectively (Supplementary Table 5). Supplementary Figure 1 shows median data completeness for each hour in a day across the analyzed biweekly segments. Similarly, we divided the actual number of GPS data points collected per hour by the expected one and computed the standard deviation of these 24 normalized values to characterize sampling constancy of the daily data. Any positive value for sampling constancy indicates fluctuations in the volume of GPS data acquired throughout a day, with greater values indicating greater fluctuations (Supplementary Table 6). Median completeness and median sampling constancy of the daily data, as computed across 14 days of a biweekly segment, were used to characterize the quality of GPS recordings acquired for that segment.

### Home stay

Home location was identified in a stepwise manner. Initially, home location was approximated by the median longitude and latitude of all GPS data points in a biweekly segment acquired between 12 am and 6 am (Chow et al. 2017; Saeb et al. 2015, 2016). To account for accidental travel and outdoor stay, all GPS data points acquired during these hours and separated by > 60 meters from the initial home location were discarded. Home location was finally determined as the median longitude and latitude of all the remaining GPS data points (Figure 1). Distance between any two GPS data points *i* and *j* was computed using the Haversine WGS84 formula as follows (Depp et al. 2019):

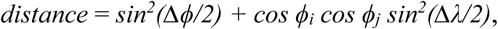

where *ϕ* and *λ* correspond to the latitude and longitude, respectively.

Home stay for a given day was specified as a ratio between the number of GPS data points separated by ≤ 60 meters from home location and the total number of GPS data points acquired on that day. Home stay values of 0 (or 0%) and 1 (or 100%) correspond to an entire day spent outside versus at home, respectively. Median home stay, as computed across 14 days of a biweekly segment, was used to characterize home stay of a study participant for that segment (Supplementary Table 7).

### Statistical analysis

A linear regression model was selected to test for a relationship between home stay and overall severity of the MDD symptoms. Specifically, home stay was used as a dependent variable, with PHQ-8 total score being used as an independent variable. Participant age at enrolment, gender (male versus female), occupational status (employed versus unemployed), median completeness and sampling constancy of the daily data in a biweekly segment were included in the model as additional explanatory variables:

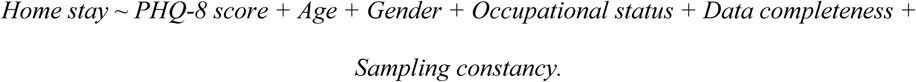

We chose home stay as a dependent variable to test its relationship not only to the severity of MDD symptoms but also to participant’s demographics and quality characteristics of the collected geolocation data in a single model in a uniform manner. For each study participant we randomly selected one of the biweekly segments generated by that participant. The model was fitted using data of the biweekly segments pooled across the participants. To obtain a confidence interval for each of the six regression coefficients, the procedure of random selection of a biweekly segment per participant followed by pooling data across the participants and fitting the model was repeated 100 times. A model parameter was deemed to be significantly related to home stay if a 95% two-sided confidence interval obtained for the regression coefficient of that parameter did not include 0. The model was fitted using data of all three sites combined (Table 2) and each clinical site separately (Supplementary Table 8).

**Table 2.** Confidence intervals and medians for the six regression coefficients of the linear regression model.

| Analyzed time frame | Statistic | Participant age | Participant gender | PHQ-8 total score | Occupational status | Median completeness of the daily data | Median sampling constancy of the daily data |
| --- | --- | --- | --- | --- | --- | --- | --- |
| Over the entire week | CI | **0.161; 0.325** | -0.272; 0.024 | **0.015; 0.184** | **-0.631; -0.279** | -0.108; 0.022 | -0.130; 0.005 |
|  | Median | **0.241** | -0.121 | **0.100** | **-0.448** | -0.044 | -0.064 |
| Weekdays only | CI | **0.188; 0.329** | -0.220; 0.058 | **0.023; 0.178** | **-0.664; -0.354** | -0.097; 0.034 | -0.110; 0.041 |
|  | Median | **0.254** | -0.061 | **0.098** | **-0.495** | -0.024 | -0.041 |
| Weekends only | CI | **0.029; 0.240** | -0.265; 0.174 | -0.079; 0.149 | **-0.535; -0.127** | -0.074; 0.101 | -0.144; 0.006 |
|  | Median | **0.148** | -0.036 | 0.052 | **-0.323** | 0.023 | -0.075 |

To test whether a relationship between home stay and the independent variables differed between weekdays and weekends, a similar approach to that above was followed. Specifically, home stay, median completeness and sampling constancy of the daily data in a biweekly segment were estimated separately for the weekdays and weekends. As a single biweekly segment included ten weekdays and only four weekend days, we used four days to generate those estimates in order to equalize variance in the estimates of weekdays and weekends. For each analyzed biweekly segment we randomly drew four weekdays 100 times. Medians of the estimates computed for each of these 100 draws were used to characterize weekdays of that segment in the model.

To account for non-normality of both dependent and independent variables as well as for differences in their variance, each variable (except for gender and occupational status) was standardized by applying the Yeo-Johnson transformation followed by the zero-mean, unit-variance normalization. All models and findings reported throughout the manuscript were obtained by using these “standardized” data. Note, however, that qualitatively similar results were obtained when using original, non-standardized data (Supplementary Table 9).

## Results

### Dataset characteristics

As of January 1, 2020, the total number of participants enrolled in the RADAR-MDD study across the three clinical sites was 432 (Supplementary Table 1). Of those, 401 (92.8%) participants had usable PHQ-8 and geolocation data (Table 1), resulting in a total of 4,273 biweekly segments generated across the sites (Supplementary Table 2). After discarding GPS data points with low accuracy (> 20 meters) and selecting only biweekly segments with 14 days of GPS recordings available, the number of biweekly segments reduced to 1,876 (43.9%; Supplementary Table 2). Imposing additional requirements on the daily median sampling period (≤ 11 minutes; Supplementary Table 3) and the daily minimum volume (≥ 48 data points; Supplementary Table 4) of the GPS data in a single biweekly segment further reduced the number of biweekly segments available for analysis to 769 (18.0%; Table 1). The latter corresponded to the data of 164 (38.0%) study participants. Table 1 lists the demographic characteristics of those participants, whereas Figure 2c shows the distribution of their ages. The majority of the study participants enrolled at each clinical site were females (range: 68.4 – 82.4%; Table 1).

**Figure 2.**
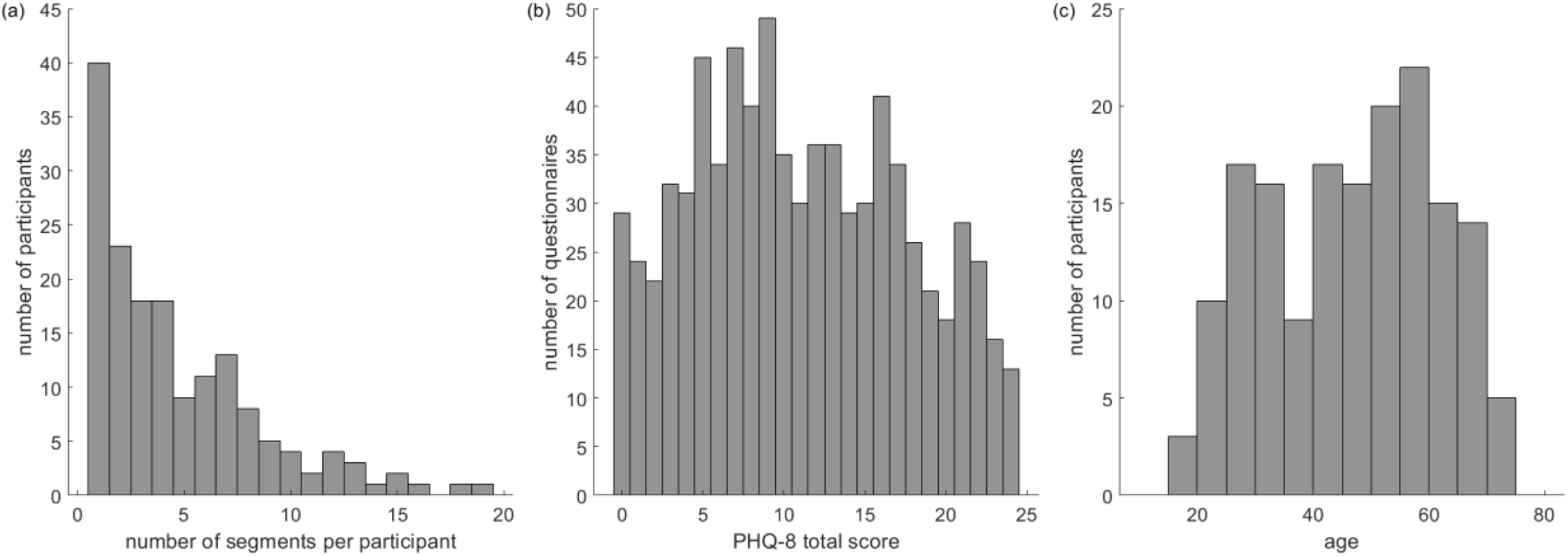
Distributions of dataset characteristics. (**a**) Number of biweekly segments available for analysis per study participant. (**b**) PHQ-8 total score. (**c**) Participant age. Data were pooled across the three clinical sites. The distributions for each individual clinical site and occupational status are presented in Supplementary Figures 2 and 3, respectively. Statistics on each presented dataset characteristic is reported in Table 1.

The number of biweekly segments available for analysis varied considerably across the sites, with VUMC (*n* = 64 segments or 8.3% of the total; Table 1) and KCL (*n* = 483 or 62.8%) providing the least and most data, respectively. The number of biweekly segments produced by a single participant varied between 1 and 19, with the median equalling 4 (25^th^-75^th^ percentiles: 2 – 7; Figure 2a). As can be seen in Figure 2b, the collected data represented all five severity categories of MDD, as specified in the PHQ-8 questionnaire (Kroenke et al. 2009), including “none-minimal” (PHQ-8 total score from 0 to 4: *n* = 138 segments or 18.0% of the total), “mild” (from 5 to 9: *n* = 214 or 27.8%%), “moderate” (from 10 to 14: *n* = 166 or 21.6%), “moderately severe” (from 15 to 19: *n* = 152 or 19.8%), and “severe” (from 20 to 24: *n* = 99 or 12.8%).

### Estimates of home stay

Over the course of the study, we can infer that participants spent most of their time at home. When no distinction between weekdays and weekends was made, median home stay across the sites was 89% (25^th^-75^th^ percentiles: 76% – 96%; Figure 3a). As expected, home stay was lower during weekdays as compared to weekends (Figure 3b, c). Specifically, median home stay across the sites was 87% (25^th^-75^th^ percentiles: 74% – 95%) and 93% (25^th^-75^th^ percentiles: 82% – 98%) when analysing data of weekdays and weekends, respectively. These observations were consistent across each clinical site (Supplementary Table 7; Supplementary Figure 4).

**Figure 3.**
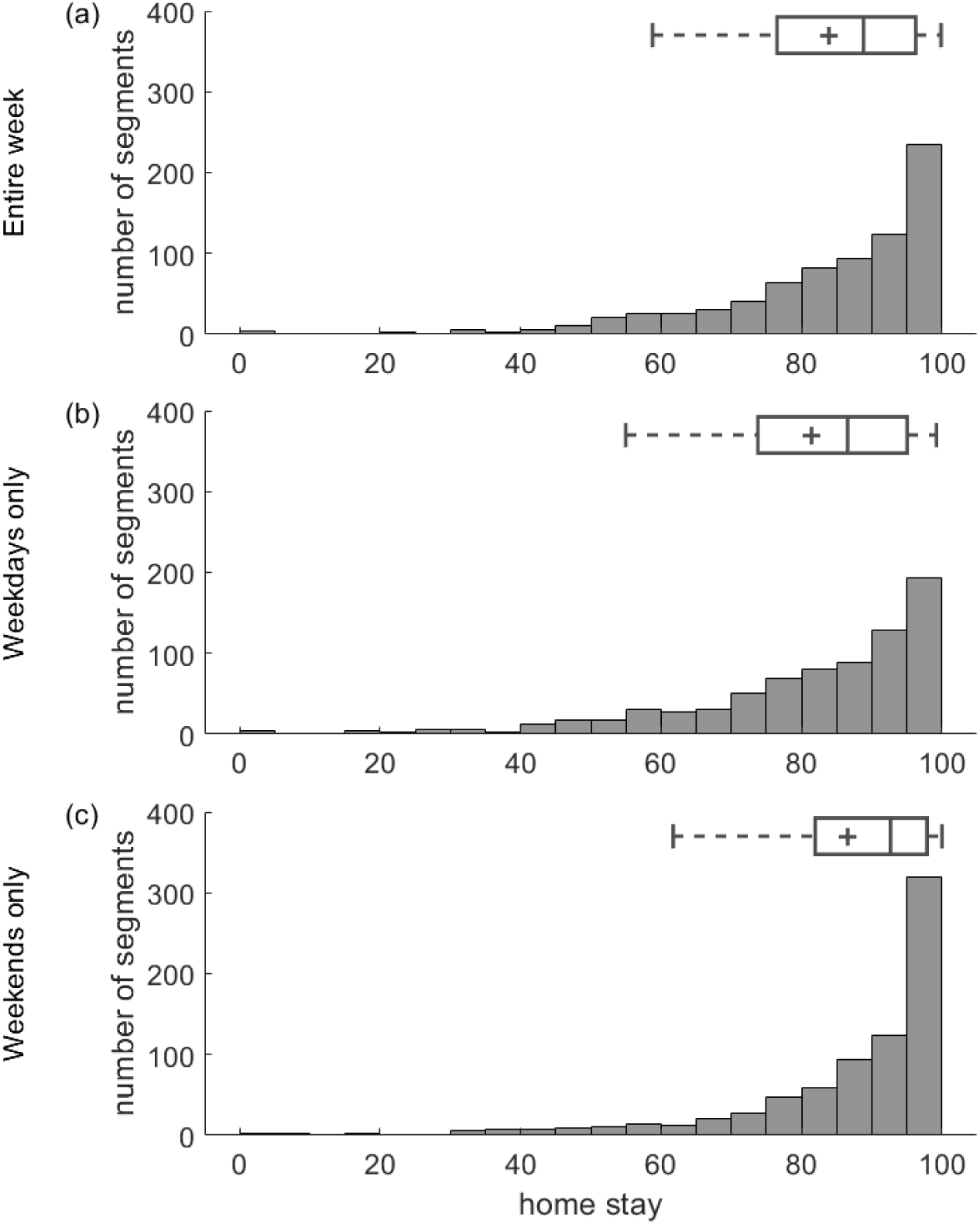
Home stay computed (**a**) over the entire week, (**b**) for weekdays and (**c**) weekends only. A grey horizontal bar and a cross in each boxplot indicate median and mean of the presented data. Data were pooled across the three clinical sites. The distributions for each individual clinical site are presented in Supplementary Figure 4.

**Figure 4.**
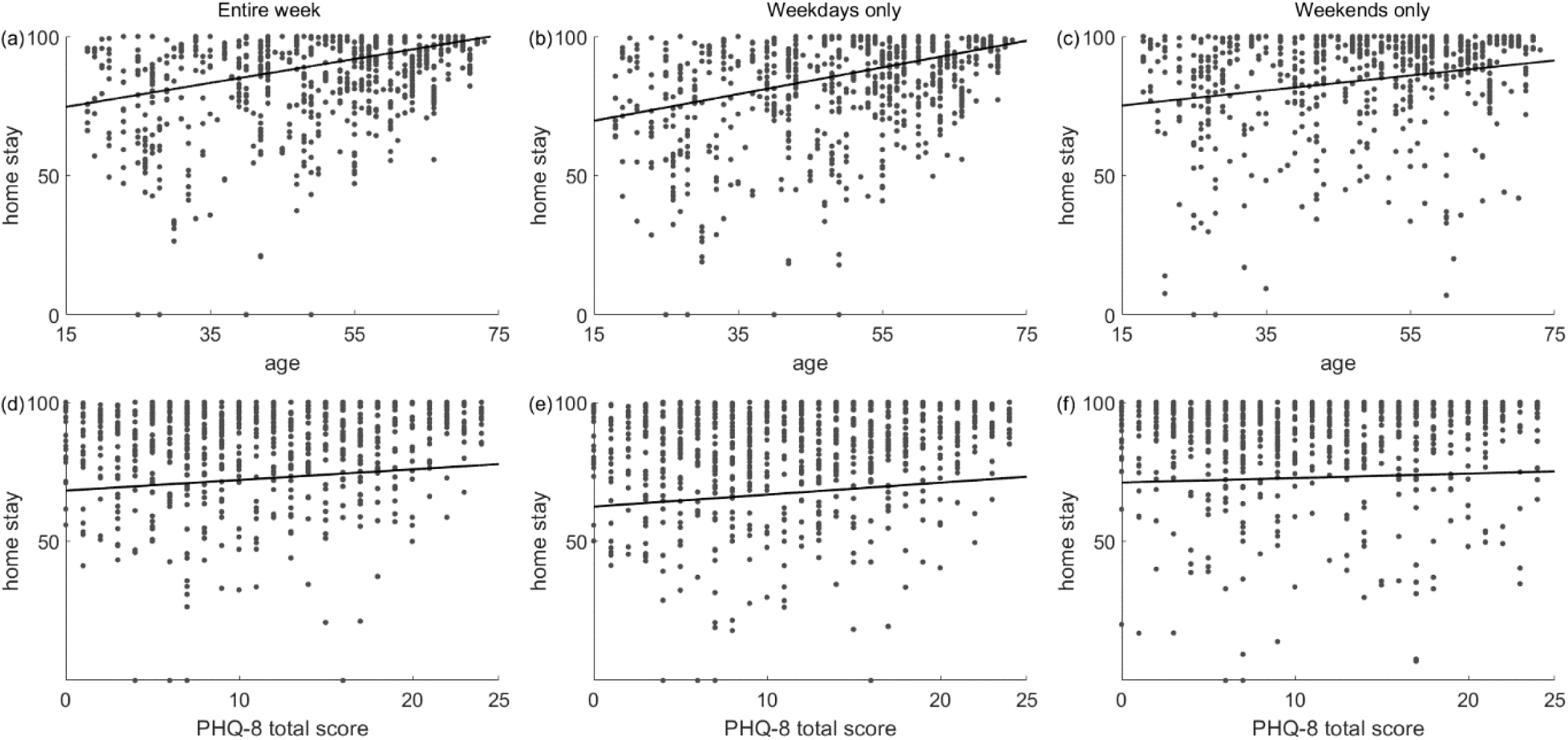
Relationship (**a, b, c**) between home stay and participant age, and (**d, e, f**) between home stay and the PHQ-8 total score, as assessed using data of (**a, d**) the entire week, (**b, e**) weekdays and (**c, f**) weekends only. Each dot indicates a single biweekly segment. Data of all biweekly segments pooled across the three clinical sites are presented. A black line in each panel corresponds to the linear fit of the presented data.

Similarly, home stay was affected by occupational status. The employed participants spent less time at home as compared to their unemployed peers. Median home stay across the sites was 82% (25^th^-75^th^ percentiles: 67% – 92%) and 94% (25^th^-75^th^ percentiles: 85% – 98%) for the employed and unemployed participants, respectively, with the difference being more prominent during weekdays (79% versus 93%) as compared to weekends (88% versus 96%). The same pattern of observations was seen across each clinical site (Supplementary Table 7; Supplementary Figure 5).

### Associations with home stay

When data were pooled across the sites and no distinction between weekdays and weekends was made, the linear regression model revealed a significant relationship between home stay and overall severity of the MDD symptoms, as captured by the PHQ-8 total score (95% two-sided confidence interval (CI) versus median: 0.015 – 0.184 versus 0.100; Table 2; Figure 4d). The latter suggests that greater overall severity of the MDD symptoms was associated with prolonged home stay. The same relationship was observed when analysing data of weekdays only (CI versus median: 0.023 – 0.178 versus 0.098; Figure 4e) but not of weekends (−0.079 – 0.149 versus 0.052; Figure 4f).

In addition, the model also revealed a significant relationship between home stay and age. Specifically, the participants spent more at home with age (CI versus median: 0.161 – 0.325 versus 0.241; Table 2; Figure 4a). Similar strength of the relationship was observed for weekdays (CI versus median: 0.188 – 0.329 versus 0.254; Figure 4b) and weekends (0.029 – 0.240 versus 0.148; Figure 4c). Furthermore, occupational status was also found to significantly modulate home stay, with the employed participants spending less time at home compared to their unemployed peers (CI versus median: −0.631 – −0.279 versus −0.448; Table 2). Similar to age, there was no significant difference in the effect of occupational status on home stay between the analyzed time frames (CI versus median, weekdays only: −0.664 – −0.354 versus −0.495; weekends only: −0.535 – −0.127 versus −0.323).

Neither gender nor median completeness and sampling constancy of the daily data in a biweekly segment had a significant impact on home stay, and this held for all the analyzed time frames (Table 2). The results of modelling for each individual clinical site obtained with standardized and original data can be found in Supplementary Tables 8 and 9, respectively.

## Discussion

Multiple studies have demonstrated associations between patterns of daily movements of an individual in an area of the primary residence and individual’s mood (Rohani et al. 2018). Here, we tested the association between home stay and overall severity of the MDD symptoms, as reflected in the PHQ-8 total score, by using data collected in the RADAR-MDD study. The participants were invited to fill in the PHQ-8 questionnaire on their mobile phones every two weeks, while the same phones were utilized to track their geographic location continuously throughout the study. We related the PHQ-8 total score, as provided by an individual, to their median daily home stay over the two weeks preceding completion of the PHQ-8 questionnaire. In addition, we investigated how the relationship between home stay and MDD symptoms severity is affected when accounting for participants age, gender, occupational status as well as for data completeness and constancy. Moreover, we also examined whether associations were stronger during the weekend or weekdays.

The participants of the RADAR-MDD study were recruited from a non-homogeneous population (i.e., clinical and community sample with a wide age range) across three clinical sites in different European countries. When we pooled the data from all sites and used the entire biweekly segment prior to the PHQ-8 completion, we found that home stay is positively associated with the PHQ-8 total score and age (Table 2). This means that a prolonged home stay relates to greater MDD symptoms severity as well as to older ages. In addition, we found that occupational status relates to home stay and unemployed people stay at home more compared to employed participants. Similar findings were observed when home stay was computed using location data from the weekdays of the biweekly segment (Table 2). Regarding the associations in weekends, we observed that age and occupational status affect the home stay but no associations with the PHQ-8 total score were found. This loss of association probably happened due to the ceiling effect (Šimkovic et al. 2019) as during the weekend the home stay values were quite large for almost all participants (Figure 3c; Supplementary Table 7). Similar findings were observed for the KCL and CIBER site (Supplementary Table 8), however no associations between home stay and PHQ-8 were found in the CIBER site because that site mainly recruited in a clinical setting, and the vast majority of the PHQ-8 values were elevated (Supplementary Figure 2b). The VUMC site comprised of only 17 participants (Table 1) whose vast majority PHQ-8 values were quite low (Supplementary Figure 2b).

Many features could be extracted from the location data of an individual. Among them are the home stay (Wahle et al. 2016), number of visited places (Saeb et al. 2016), location entropy (i.e., quantifies uniformity in the distribution of the time that is spent across different visited places) (Saeb et al. 2015, 2016), maximal distance from home and total distance travelled (Depp et al. 2019). Notably, several studies that investigated the relationship between mental health disorders and mobility patterns have focused on home stay features (Chow et al. 2017; Depp et al. 2019; Tung et al. 2016). In this study, we also used home stay to quantify the mobility behavior of the participants as clinicians consider it an important social disengagement indicator (Chow et al. 2017). Moreover, it has been demonstrated that home stay has a strong negative relationship with location entropy (Rohani et al. 2018 and references therein). In addition, features that quantify the distance between location places (e.g., total distance travelled or maximal distance from home) were not so relevant in our dataset since the RADAR participants lived in urban or rural places and, therefore, the notion of distance was not informative.

The finding that home stay has a positive relationship with MDD severity symptoms has been documented in several previous studies (Rohani et al. 2018; Saeb et al. 2015, 2016; Wahle et al. 2016). However, to the best of our knowledge there is no study that encompasses data from multiple sites which were collected from a non-homogeneous population with confirmed clinical diagnosis of MDD and accounts for participants’ age, occupational status and data quality. Several previous studies used a homogeneous dataset with regard to the age (i.e., students’ population) (Chow et al. 2017; Farhan et al. 2016) whereas, the age range of the RADAR participants was from 18 to 73 years (Figure 2c). The finding that home stay associates positively with age indicates that the participants age could have an effect on the strength of the relationship between home stay and severity of MDD symptoms. Such relationship is expected to be stronger when the study participants are young and weaker for old participants, since old people tend to stay at home more. In addition, all RADAR-MDD participants had a clinical diagnosis of MDD and many of them manifested severe symptoms of MDD as indicated from the high PHQ-8 total scores (Figure 1b), whereas participants from previous studies did not undergo clinical interviews and had overall low depression scores (Canzian 2015; Saeb et al. 2015; Saeb et al. 2016; Chow et al. 2017).

It has been reported (Depp et al. 2019; Rohani et al. 2018) that there is no standardized protocol regarding preprocessing of location data from smartphones and therefore important preprocessing steps such as selection of accuracy error threshold as well as missing rate of data could have a big impact in the reported results. In this study we did not use all location data from the RADAR-MDD participants, but we applied stringent selection criteria (i.e. presence of daily recordings thought the biweekly segment; maximum median daily sampling period of 11min; accuracy error of the GPS signal less than 20m) in order to ensure best quality of the data. In addition, we presented in detail the information for all collected and analysed data (Supplementary Tables 1,2, 3 and 4).

Some participants in the RADAR-MDD study were following antidepressant treatment and some of them reported comorbidity with physical illness e.g. fybromyalgia. Additionally, there were participants that were off-sick or reported ill health. Antidepressants may cause a wide range of side effects, including headaches, fatigue, weight gain, drowsiness, and dizziness (Ferguson 2001; Wang et al. 2018). Individuals that experience any or all of these side effects or have comorbidity with physical illness are likely to spend more time at home than outdoors. This could have inflated the reported estimates of home stay (Figure 3) and, thus, distorted the strength of the relationship between home stay and overall severity of the MDD symptoms. The effect of medication and physical comorbidity on the relationship between daily mobility patterns and severity of MDD symptoms was beyond the scope of this work, though it certainly warrants further research.

In conclusion, in individuals with a diagnosis of MDD we show that those with a higher level of home stay during a 2-week ambulatory assessment period show higher depression severity. Our findings illustrate that passive sensing of individuals with depression is feasible and could provide clinically useful information to monitor the course of illness in patients with MDD. Whether the findings are causally associated with changes in depressive symptoms or represent behavioral manifestations of depression is unclear. Future analyses will tease out whether home stay predicts future depressive relapse.

## Data Availability

The data can be requested from the RADAR-MDD consortium

## Code and Data Availability

The code and data can be requested from the RADAR-MDD consortium.

## Acknowledgements

The RADAR-CNS project has received funding from the Innovative Medicines Initiative (IMI) 2 Joint Undertaking under grant agreement no. 115902. This Joint Undertaking receives support from the European Union’s Horizon 2020 research and innovation programme and EFPIA (www.imi.europa.eu). This communication reflects the views of the RADAR-CNS consortium and neither IMI nor the European Union and EFPIA are liable for any use that may be made of the information contained herein.

Participant recruitment in Amsterdam was partially accomplished through Hersenonderzoek.nl, a Dutch online registry that facilitates participant recruitment for neuroscience studies (www.hersenonderzoek.nl). Hersenonderzoek.nl is funded by ZonMw-Memorabel (project no. 73305095003), a project in the context of the Dutch Deltaplan Dementie, Gieskes-Strijbis Foundation, the Alzheimer’s Society in the Netherlands and Brain Foundation Netherlands. Participants from the CIBER group were recruited from the following Heath Care centers: Parc Sanitari Sant Joan de Déu Network services, Institut Català de la Salut-IDIAP Jordi Gol, (Catalonia) Institut Pere Mata Hospital Clínico San Carlos, Madrid, Spain. We also acknowledge the great contribution of all participants with MDD across sites.

This paper represents independent research part-funded by the National Institute for Health Research (NIHR) Biomedical Research Centre at South London and Maudsley NHS Foundation Trust and King’s College London. The views expressed are those of the authors and not necessarily those of the NHS, the NIHR or the Department of Health and Social Care.

