## Supplementary Material for "Home stay reflects symptoms severity in major depressive disorder: A multicenter observational study using geolocation data from smartphones"

**Affiliations:**

King’s College London

**Running title:** Home stay reflects symptom severity in MDD.

**Supplementary Information**

**Supplementary Table 1.** Total number of participants enrolled at each clinical site.

**Supplementary Table 2.** Number of the biweekly segments available for analysis as a function of accuracy of the GPS signal.

**Supplementary Table 3.** Number of the days with GPS recordings available for analysis as a function of daily median sampling period of the GPS signal.

**Supplementary Table 4.** Number of the biweekly segments available for analysis as a function of daily number of acquired GPS data points.

**Supplementary Table 5.** Statistics on median *completeness* of the daily data in biweekly segments for each clinical site, analyzed time frame and occupational status.

**Supplementary Table 6.** Statistics on median *sampling constancy* of the daily data in biweekly segments for each clinical site, analyzed time frame and occupational status.

**Supplementary Table 7.** Statistics on home stay for each clinical site, analyzed time frame and occupational status.

**Supplementary Table 8.** Confidence intervals and medians for the six regression coefficients of the linear regression model obtained with *standardized* data of each clinical site separately.

**Supplementary Table 9.** Confidence intervals and medians for the six regression coefficients of the linear regression model obtained with *original*, *non-standardized* data of each clinical site separately and all three sites combined.

**Supplementary Figure 1.** Completeness of the analyzed geolocation data for each hour in a day.

**Supplementary Figure 2.** Distributions of dataset characteristics for each clinical site separately.

**Supplementary Figure 3.** Distributions of dataset characteristics for the employed versus unemployed participants.

**Supplementary Figure 4.** Home stay computed (**a**) over the entire week, (**b**) for weekdays and (**c**) weekends only for each clinical site separately.

**Supplementary Figure 5.** Distributions of home stay for the employed versus unemployed participants.

**Supplementary Table 1.** Total number of participants enrolled at each clinical site.

| Characteristic | Participant gender (% in a group) | | |
| --- | --- | --- | --- |
|  | Male | Female | Both genders |
| Clinical site: KCL | | | |
| Number of participants | 64 (24.9%) | 193 (75.1%) | **257 (100.0%)** |
| with at least one PHQ-8 filled in | 56 (21.8%) | 181 (70.4%) | 237 (92.2%) |
| with at least one GPS data point collected | 62 (24.1%) | 187 (72.8%) | 249 (96.9%) |
| with both PHQ-8 and GPS data collected | 55 (21.4%) | 177 (68.9%) | 232 (90.3%) |
| Clinical site: CIBER | | | |
| Number of participants | 34 (27.9%) | 88 (72.1%) | **122 (100.0%)** |
| with at least one PHQ-8 filled in | 32 (26.2%) | 85 (69.7%) | 117 (95.9%) |
| with at least one GPS data point collected | 34 (27.9%) | 85 (69.7%) | 119 (97.6%) |
| with both PHQ-8 and GPS data collected | 32 (26.2%) | 84 (68.9%) | 116 (95.1%) |
| Clinical site: VUMC | | | |
| Number of participants | 12 (22.6%) | 41 (77.4%) | **53 (100.0%)** |
| with at least one PHQ-8 filled in | 12 (22.6%) | 41 (77.4%) | 53 (100.0%) |
| with at least one GPS data point collected | 12 (22.6%) | 41 (77.4%) | 53 (100.0%) |
| with both PHQ-8 and GPS data collected | 12 (22.6%) | 41 (77.4%) | 53 (100.0%) |

The number of participants for each selection criterion was normalized by the total number of participants enrolled at a site (highlighted in bold), with the resulting percentage being indicated in parentheses.

**Supplementary Table 2.** Number of the biweekly segments available for analysis as a function of accuracy of the GPS signal.

| Clinical site | Statistic | Accuracy of the GPS signal (meters) | | | | | Total number of the biweekly segments |
| --- | --- | --- | --- | --- | --- | --- | --- |
|  |  | ≤ 10 | ≤ 15 | ≤ 20 | ≤ 25 | ≤ 30 |  |
| KCL | n  % | 516  20.1% | 740  28.8% | **1247 48.5%** | 1280 49.8% | 1284 49.9% | 2571  100.0% |
| CIBER | n  % | 97  7.6% | 177  14.0% | **439**  **34.6%** | 444  35.0% | 444  35.0% | 1268  100.0% |
| VUMC | n  % | 51  11.8% | 83  19.1% | **190**  **43.8%** | 193  44.5% | 194  44.7% | 434  100.0% |
| All sites | n  % | 664  15.5% | 1000  23.4% | **1876**  **43.9%** | 1917  44.9% | 1922  45.0% | 4273  100.0% |

All GPS data points with a lower accuracy than specified were discarded. Only biweekly segments with 14 days of GPS recordings available prior to completion of a PHQ-8 questionnaire and with each day having at least two GPS data points collected were analyzed. n indicates the number of the biweekly segments that meet the selection criteria and can be analyzed for a given accuracy of the GPS signal, whereas % indicates their percentage relative to the total number of the biweekly segments obtained at a site. The latter correspond to the biweekly segments generated regardless of the GPS signal accuracy, the number of days with GPS recordings available, and the number of GPS data points collected per day of recordings. “All sites” denotes data pooled across the three sites.

**Supplementary Table 3.** Number of the days with GPS recordings available for analysis as a function of daily median sampling period of the GPS signal.

| Daily median sampling period (minutes) | Clinical site | | | |
| --- | --- | --- | --- | --- |
|  | KCL | CIBER | VUMC | All sites |
| ≤ 5.0 | 499 (2.9%) | 287 (4.7%) | 59 (2.2%) | 845 (3.2%) |
| ≤ 6.0 | 8219 (47.1%) | 3899 (63.4%) | 1439 (54.1%) | 13557 (51.6%) |
| ≤ 7.0 | 10014 (57.4%) | 4403 (71.6%) | 1596 (60.0%) | 16013 (61.0%) |
| ≤ 8.0 | 10885 (62.3%) | 4581 (74.5%) | 1665 (62.6%) | 17131 (65.2%) |
| ≤ 9.0 | 11699 (67.0%) | 4726 (76.9%) | 1739 (65.4%) | 18164 (69.2%) |
| ≤ 10.0 | 12990 (74.4%) | 4921 (80.1%) | 1884 (70.8%) | 19795 (75.4%) |
| ≤ 11.0 | 13996 (80.2%) | 5078 (82.6%) | 1985 (74.6%) | 21059 (80.2%) |
| ≤ 12.0 | 14226 (81.5%) | 5130 (83.5%) | 2014 (75.7%) | 21370 (81.4%) |
| ≤ 13.0 | 14419 (82.6%) | 5164 (84.0%) | 2033 (76.4%) | 21616 (82.3%) |
| ≤ 14.0 | 14589 (83.6%) | 5201 (84.6%) | 2054 (77.2%) | 21844 (83.2%) |
| ≤ 15.0 | 14944 (85.6%) | 5406 (88.0%) | 2170 (81.6%) | 22520 (85.7%) |
| # Days in total | 17458 (100.0%) | 6146 (100.0%) | 2660 (100.0%) | 26264 (100.0%) |
| # Segments | **1247 (= 17458 / 14)** | **439 (= 6146 / 14)** | **190 (= 2660 / 14)** | **1876 = (26264 / 14)** |

Only biweekly segments with 14 days of GPS recordings available prior to completion of a PHQ-8 questionnaire, with all GPS data points in a segment having accuracy ≤ 20 meters and with each day having at least two GPS data points collected were analyzed (highlighted in bold in Supplementary Tables 2 and 3). The number of days for each sampling period was normalized by the total number of days with GPS recordings obtained at a site, with the resulting percentage being indicated in parentheses. “All sites” denotes data pooled across the three sites. Abbreviations: # – number of.

**Supplementary Table 4.** Number of the biweekly segments available for analysis as a function of daily number of acquired GPS data points.

| Daily number of acquired GPS data points | Corresponding sampling period (minutes) | Clinical site | | | |
| --- | --- | --- | --- | --- | --- |
|  |  | KCL | CIBER | VUMC | All sites |
| ≥ 144 | ≤ 10 | 53 | 30 | 6 | 89 |
| ≥ 131 | ≤ 11 | 84 | 41 | 17 | 142 |
| ≥ 96 | ≤ 15 | 244 | 104 | 34 | 382 |
| ≥ 72 | ≤ 20 | 379 | 167 | 47 | 593 |
| ≥ 48 | ≤ 30 | 483 | 222 | 64 | 769 |

Only biweekly segments with 14 days of GPS recordings available prior to completion of a PHQ-8 questionnaire, with all GPS data points in a segment having accuracy ≤ 20 meters, with each day having at least two GPS data points collected and daily median sampling period ≤ 11 minutes were analyzed. Note that although two independent days of GPS recordings may have the same median sampling period, they still can differ in the number of acquired GPS data points. “All sites” denotes data pooled across the three sites.

**Supplementary Table 5.** Statistics on median *completeness* of the daily data in biweekly segments for each clinical site, analyzed time frame and occupational status.

| Clinical site | Analyzed time frame | Occupational status | # Biweekly segments | 25^th^ percentile | Median | 75^th^ percentile |
| --- | --- | --- | --- | --- | --- | --- |
| KCL | Over the entire week | Employed | 277 | 0.46 | 0.55 | 0.65 |
|  |  | Unemployed | 204 | 0.43 | 0.55 | 0.64 |
|  |  | All combined | 481 | 0.44 | 0.55 | 0.65 |
|  | Weekdays only | Employed | 277 | 0.46 | 0.55 | 0.64 |
|  |  | Unemployed | 204 | 0.43 | 0.55 | 0.65 |
|  |  | All combined | 481 | 0.44 | 0.55 | 0.64 |
|  | Weekends only | Employed | 277 | 0.44 | 0.55 | 0.64 |
|  |  | Unemployed | 204 | 0.42 | 0.52 | 0.62 |
|  |  | All combined | 481 | 0.42 | 0.53 | 0.63 |
| CIBER | Over the entire week | Employed | 64 | 0.49 | 0.56 | 0.62 |
|  |  | Unemployed | 158 | 0.41 | 0.51 | 0.64 |
|  |  | All combined | 222 | 0.42 | 0.53 | 0.63 |
|  | Weekdays only | Employed | 64 | 0.5 | 0.57 | 0.63 |
|  |  | Unemployed | 158 | 0.42 | 0.52 | 0.63 |
|  |  | All combined | 222 | 0.44 | 0.54 | 0.63 |
|  | Weekends only | Employed | 64 | 0.46 | 0.55 | 0.61 |
|  |  | Unemployed | 158 | 0.38 | 0.48 | 0.64 |
|  |  | All combined | 222 | 0.41 | 0.52 | 0.63 |
| VUMC | Over the entire week | Employed | 41 | 0.44 | 0.56 | 0.61 |
|  |  | Unemployed | 23 | 0.51 | 0.54 | 0.59 |
|  |  | All combined | 64 | 0.48 | 0.55 | 0.6 |
|  | Weekdays only | Employed | 41 | 0.45 | 0.53 | 0.59 |
|  |  | Unemployed | 23 | 0.48 | 0.55 | 0.58 |
|  |  | All combined | 64 | 0.46 | 0.54 | 0.59 |

Continued on the next page.

| Clinical site | Analyzed time frame | Occupational status | # Biweekly segments | 25^th^ percentile | Median | 75^th^ percentile |
| --- | --- | --- | --- | --- | --- | --- |
| VUMC | Weekends only | Employed | 41 | 0.48 | 0.55 | 0.64 |
|  |  | Unemployed | 23 | 0.52 | 0.57 | 0.62 |
|  |  | All combined | 64 | 0.51 | 0.56 | 0.63 |
| All sites | Over the entire week | Employed | 382 | 0.47 | 0.55 | 0.64 |
|  |  | Unemployed | 385 | 0.42 | 0.53 | 0.64 |
|  |  | All combined | 767 | 0.44 | 0.54 | 0.64 |
|  | Weekdays only | Employed | 382 | 0.46 | 0.55 | 0.64 |
|  |  | Unemployed | 385 | 0.43 | 0.54 | 0.64 |
|  |  | All combined | 767 | 0.44 | 0.55 | 0.64 |
|  | Weekends only | Employed | 382 | 0.44 | 0.55 | 0.63 |
|  |  | Unemployed | 385 | 0.41 | 0.52 | 0.63 |
|  |  | All combined | 767 | 0.42 | 0.53 | 0.63 |

“All sites” denotes data pooled across the three sites. “All combined” denotes data of both employed and unemployed participants pooled together. Note that two biweekly segments obtained at KCL were discarded due to missing data on the occupational status. Abbreviations: # – number of.

**Supplementary Table 6.** Statistics on median *sampling constancy* of the daily data in biweekly segments for each clinical site, analyzed time frame and occupational status.

| Clinical site | Analyzed time frame | Occupational status | # Biweekly segments | 25^th^ percentile | Median | 75^th^ percentile |
| --- | --- | --- | --- | --- | --- | --- |
| KCL | Over the entire week | Employed | 277 | 0.26 | 0.31 | 0.35 |
|  |  | Unemployed | 204 | 0.24 | 0.29 | 0.32 |
|  |  | All combined | 481 | 0.25 | 0.3 | 0.34 |
|  | Weekdays only | Employed | 277 | 0.26 | 0.3 | 0.35 |
|  |  | Unemployed | 204 | 0.24 | 0.28 | 0.32 |
|  |  | All combined | 481 | 0.25 | 0.29 | 0.34 |
|  | Weekends only | Employed | 277 | 0.26 | 0.31 | 0.36 |
|  |  | Unemployed | 204 | 0.24 | 0.29 | 0.33 |
|  |  | All combined | 481 | 0.25 | 0.3 | 0.35 |
| CIBER | Over the entire week | Employed | 64 | 0.24 | 0.27 | 0.3 |
|  |  | Unemployed | 158 | 0.27 | 0.31 | 0.35 |
|  |  | All combined | 222 | 0.26 | 0.3 | 0.34 |
|  | Weekdays only | Employed | 64 | 0.24 | 0.27 | 0.3 |
|  |  | Unemployed | 158 | 0.27 | 0.31 | 0.34 |
|  |  | All combined | 222 | 0.26 | 0.29 | 0.34 |
|  | Weekends only | Employed | 64 | 0.25 | 0.29 | 0.31 |
|  |  | Unemployed | 158 | 0.26 | 0.31 | 0.36 |
|  |  | All combined | 222 | 0.26 | 0.3 | 0.35 |
| VUMC | Over the entire week | Employed | 41 | 0.28 | 0.3 | 0.33 |
|  |  | Unemployed | 23 | 0.25 | 0.26 | 0.31 |
|  |  | All combined | 64 | 0.26 | 0.29 | 0.33 |
|  | Weekdays only | Employed | 41 | 0.27 | 0.3 | 0.33 |
|  |  | Unemployed | 23 | 0.24 | 0.26 | 0.32 |
|  |  | All combined | 64 | 0.26 | 0.29 | 0.32 |

Continued on the next page.

| Clinical site | Analyzed time frame | Occupational status | # Biweekly segments | 25^th^ percentile | Median | 75^th^ percentile |
| --- | --- | --- | --- | --- | --- | --- |
| VUMC | Weekends only | Employed | 41 | 0.26 | 0.29 | 0.33 |
|  |  | Unemployed | 23 | 0.25 | 0.27 | 0.29 |
|  |  | All combined | 64 | 0.26 | 0.28 | 0.33 |
| All sites | Over the entire week | Employed | 382 | 0.26 | 0.3 | 0.34 |
|  |  | Unemployed | 385 | 0.25 | 0.3 | 0.33 |
|  |  | All combined | 767 | 0.26 | 0.3 | 0.34 |
|  | Weekdays only | Employed | 382 | 0.26 | 0.3 | 0.34 |
|  |  | Unemployed | 385 | 0.25 | 0.29 | 0.33 |
|  |  | All combined | 767 | 0.25 | 0.29 | 0.34 |
|  | Weekends only | Employed | 382 | 0.26 | 0.3 | 0.35 |
|  |  | Unemployed | 385 | 0.25 | 0.3 | 0.34 |
|  |  | All combined | 767 | 0.25 | 0.3 | 0.35 |

“All sites” denotes data pooled across the three sites. “All combined” denotes data of both employed and unemployed participants pooled together. Note that two biweekly segments obtained at KCL were discarded due to missing data on the occupational status. Abbreviations: # – number of.

**Supplementary Table 7.** Statistics on home stay for each clinical site, analyzed time frame and occupational status.

| Clinical site | Analyzed time frame | Occupational status | # Biweekly segments | 25^th^ percentile | Median | 75^th^ percentile |
| --- | --- | --- | --- | --- | --- | --- |
| KCL | Over the entire week | Employed | 277 | 66% | 81% | 94% |
|  |  | Unemployed | 204 | 85% | 94% | 98% |
|  |  | All combined | 481 | 74% | 89% | 96% |
|  | Weekdays only | Employed | 277 | 62% | 78% | 91% |
|  |  | Unemployed | 204 | 82% | 93% | 98% |
|  |  | All combined | 481 | 71% | 86% | 95% |
|  | Weekends only | Employed | 277 | 74% | 89% | 96% |
|  |  | Unemployed | 204 | 89% | 97% | 99% |
|  |  | All combined | 481 | 81% | 93% | 98% |
| CIBER | Over the entire week | Employed | 64 | 71% | 81% | 87% |
|  |  | Unemployed | 158 | 86% | 92% | 99% |
|  |  | All combined | 222 | 81% | 88% | 97% |
|  | Weekdays only | Employed | 64 | 62% | 81% | 87% |
|  |  | Unemployed | 158 | 83% | 92% | 98% |
|  |  | All combined | 222 | 78% | 87% | 96% |
|  | Weekends only | Employed | 64 | 78% | 86% | 94% |
|  |  | Unemployed | 158 | 87% | 95% | 99% |
|  |  | All combined | 222 | 83% | 93% | 98% |
| VUMC | Over the entire week | Employed | 41 | 72% | 84% | 91% |
|  |  | Unemployed | 23 | 80% | 97% | 98% |
|  |  | All combined | 64 | 77% | 87% | 97% |
|  | Weekdays only | Employed | 41 | 69% | 81% | 89% |
|  |  | Unemployed | 23 | 77% | 97% | 98% |
|  |  | All combined | 64 | 73% | 84% | 96% |

Continued on the next page.

| Clinical site | Analyzed time frame | Occupational status | # Biweekly segments | 25^th^ percentile | Median | 75^th^ percentile |
| --- | --- | --- | --- | --- | --- | --- |
| VUMC | Weekends only | Employed | 41 | 80% | 88% | 93% |
|  |  | Unemployed | 23 | 89% | 97% | 98% |
|  |  | All combined | 64 | 81% | 91% | 97% |
| All sites | Over the entire week | Employed | 382 | 67% | 82% | 92% |
|  |  | Unemployed | 385 | 85% | 94% | 98% |
|  |  | All combined | 767 | 76% | 89% | 96% |
|  | Weekdays only | Employed | 382 | 63% | 79% | 90% |
|  |  | Unemployed | 385 | 82% | 93% | 98% |
|  |  | All combined | 767 | 74% | 87% | 95% |
|  | Weekends only | Employed | 382 | 76% | 88% | 96% |
|  |  | Unemployed | 385 | 88% | 96% | 99% |
|  |  | All combined | 767 | 82% | 93% | 98% |

“All sites” denotes data pooled across the three sites. “All combined” denotes data of both employed and unemployed participants pooled together. Note that two biweekly segments obtained at KCL were discarded due to missing data on the occupational status. Abbreviations: # – number of.

**Supplementary Table 8.** Confidence intervals and medians for the six regression coefficients of the linear regression model obtained with *standardized* data of each clinical site separately.

| Clinical site | Statistic | Participant age | Participant gender | PHQ-8 total score | Occupational status | Median completeness of the daily data | Median sampling constancy of the daily data |
| --- | --- | --- | --- | --- | --- | --- | --- |
| Over the entire week | | | | | | | |
| KCL | CI | **0.169; 0.400** | **-0.405; -0.053** | **0.045; 0.308** | **-0.671; -0.237** | -0.150; 0.029 | -0.172; 0.063 |
|  | Median | **0.297** | **-0.233** | **0.196** | **-0.447** | -0.059 | -0.088 |
| CIBER | CI | **0.024; 0.372** | -0.327; 0.138 | -0.155; 0.109 | **-0.951; -0.396** | -0.142; 0.088 | -0.002; 0.186 |
|  | Median | **0.213** | -0.074 | -0.021 | **-0.713** | -0.011 | 0.075 |
| VUMC | CI | -0.054; 0.299 | **0.401; 1.112** | -0.544; 0.095 | -0.583; 0.414 | -0.006; 0.387 | -0.300; 0.199 |
|  | Median | 0.097 | **0.736** | -0.218 | -0.133 | 0.189 | -0.047 |
| Weekdays only | | | | | | | |
| KCL | CI | **0.195; 0.396** | -0.303; 0.019 | **0.060; 0.269** | **-0.726; -0.280** | -0.146; 0.043 | -0.182; 0.012 |
|  | Median | **0.295** | -0.172 | **0.162** | **-0.535** | -0.038 | -0.063 |
| CIBER | CI | **0.036; 0.401** | -0.273; 0.288 | -0.172; 0.132 | **-0.977; -0.388** | -0.151; 0.073 | -0.021; 0.274 |
|  | Median | **0.210** | 0.028 | 0.008 | **-0.673** | -0.021 | 0.100 |

Continued on the next page.

| Clinical site | Statistic | Participant age | Participant gender | PHQ-8 total score | Occupational status | Median completeness of the daily data | Median sampling constancy of the daily data |
| --- | --- | --- | --- | --- | --- | --- | --- |
| Weekdays only | | | | | | | |
| VUMC | CI | **0.025; 0.405** | **0.445; 1.081** | -0.563; 0.157 | -0.357; 0.370 | **0.123; 0.477** | -0.286; 0.284 |
|  | Median | **0.179** | **0.723** | -0.236 | -0.090 | **0.332** | -0.003 |
| Weekends only | | | | | | | |
| KCL | CI | **0.075; 0.320** | -0.377; 0.156 | -0.038; 0.229 | **-0.630; -0.079** | -0.059; 0.148 | -0.186; 0.074 |
|  | Median | **0.211** | -0.109 | 0.080 | **-0.344** | 0.039 | -0.070 |
| CIBER | CI | -0.202; 0.296 | -0.595; 0.172 | -0.267; 0.214 | -0.730; 0.141 | -0.125; 0.146 | -0.199; 0.136 |
|  | Median | 0.004 | -0.113 | -0.024 | -0.320 | 0.012 | -0.031 |
| VUMC | CI | -0.201; 0.330 | **0.316; 1.306** | -0.370; 0.466 | -1.101; 0.453 | -0.342; 0.234 | -0.445; 0.060 |
|  | Median | 0.064 | **0.728** | 0.012 | -0.307 | -0.079 | -0.172 |

The positive sign of the regression coefficients that correspond to the categorical variables, i.e. gender and occupational status, indicate larger home stay for male and employed as compared to female and unemployed participants, respectively. All reported confidence intervals are 95% two-sided intervals. Confidence intervals that do not include 0 are highlighted in bold. Abbreviations: CI – confidence interval.

**Supplementary Table 9.** Confidence intervals and medians for the six regression coefficients of the linear regression model obtained with *original*, *non-standardized* data of each clinical site separately and all three sites combined.

| Clinical site | Statistic | Participant age | Participant gender | PHQ-8 total score | Occupational status | Median completeness of the daily data | Median sampling constancy of the daily data |
| --- | --- | --- | --- | --- | --- | --- | --- |
| Over the entire week | | | | | | | |
| KCL | CI | **0.002; 0.005** | **-0.096; -0.012** | **0.000; 0.008** | **-0.119; -0.033** | -0.176; 0.063 | **-0.412; -0.098** |
|  | Median | **0.004** | **-0.054** | **0.005** | **-0.069** | -0.060 | **-0.263** |
| CIBER | CI | **0.001; 0.005** | -0.056; 0.009 | -0.004; 0.001 | **-0.147; -0.064** | -0.106; 0.113 | **0.022; 0.442** |
|  | Median | **0.003** | -0.018 | -0.001 | **-0.099** | 0.011 | **0.242** |
| VUMC | CI | **0.000; 0.006** | **0.025; 0.134** | -0.011; 0.012 | -0.082; 0.277 | **0.013; 0.734** | -1.667; 0.468 |
|  | Median | **0.002** | **0.085** | -0.003 | -0.029 | **0.169** | -0.112 |
| All sites | CI | **0.002; 0.004** | **-0.067; -0.005** | **0.000; 0.005** | **-0.102; -0.030** | -0.114; 0.054 | **-0.360; -0.067** |
|  | Median | **0.003** | **-0.034** | **0.002** | **-0.069** | -0.037 | **-0.210** |
| Weekdays only | | | | | | | |
| KCL | CI | **0.002; 0.006** | **-0.103; -0.002** | **0.001; 0.008** | **-0.143; -0.055** | -0.195; 0.033 | **-0.414; -0.046** |
|  | Median | **0.004** | **-0.053** | **0.005** | **-0.103** | -0.082 | **-0.251** |

Continued on the next page.

| Clinical site | Statistic | Participant age | Participant gender | PHQ-8 total score | Occupational status | Median completeness of the daily data | Median sampling constancy of the daily data |
| --- | --- | --- | --- | --- | --- | --- | --- |
| Weekdays only | | | | | | | |
| CIBER | CI | **0.001; 0.005** | -0.075; 0.042 | -0.004; 0.002 | **-0.155; -0.050** | -0.164; 0.098 | -0.130; 0.588 |
|  | Median | **0.003** | -0.009 | -0.001 | **-0.112** | 0.010 | 0.265 |
| VUMC | CI | **0.000; 0.007** | **0.019; 0.148** | -0.013; 0.010 | -0.074; 0.281 | **0.100; 0.871** | -1.679; 0.596 |
|  | Median | **0.002** | **0.087** | -0.004 | -0.014 | **0.321** | -0.056 |
| All sites | CI | **0.002; 0.005** | -0.057; 0.001 | **0.000; 0.005** | **-0.125; -0.045** | -0.133; 0.031 | **-0.373; -0.055** |
|  | Median | **0.004** | -0.029 | **0.003** | **-0.085** | -0.057 | **-0.185** |
| Weekends only | | | | | | | |
| KCL | CI | **0.001; 0.004** | -0.087; 0.029 | -0.001; 0.006 | -0.083; 0.006 | -0.030; 0.237 | -0.338; 0.032 |
|  | Median | **0.003** | -0.028 | 0.002 | -0.045 | 0.104 | -0.151 |
| CIBER | CI | -0.002; 0.003 | -0.082; 0.048 | -0.007; 0.002 | -0.083; 0.026 | -0.131; 0.189 | -0.316; 0.495 |
|  | Median | 0.000 | -0.012 | -0.003 | -0.024 | 0.022 | 0.033 |
| VUMC | CI | -0.001; 0.005 | **0.007; 0.125** | -0.007; 0.014 | -0.108; 0.195 | -0.377; 0.192 | -1.026; 0.128 |
|  | Median | 0.001 | **0.067** | 0.001 | -0.027 | -0.014 | -0.281 |

Continued on the next page.

| Clinical site | Statistic | Participant age | Participant gender | PHQ-8 total score | Occupational status | Median completeness of the daily data | Median sampling constancy of the daily data |
| --- | --- | --- | --- | --- | --- | --- | --- |
| Weekends only | | | | | | | |
| All sites | CI | **0.001; 0.003** | -0.062; 0.020 | -0.002; 0.004 | -0.073; 0.004 | -0.023; 0.153 | -0.269; 0.063 |
|  | Median | **0.002** | -0.015 | 0.001 | -0.031 | 0.070 | -0.124 |

“All sites” denotes data pooled across the three sites. The positive sign of the regression coefficients that correspond to the categorical variables, i.e. gender and occupational status, indicate larger home stay for male and employed as compared to female and unemployed participants, respectively. Note that the regression coefficients indicate changes in proportion but not in percentage of home stay (e.g., one unit increase in independent variable *X* with the regression coefficient of 0.13 will increase home stay by 0.13 that corresponds to 13%). All reported confidence intervals are 95% two-sided intervals. Confidence intervals that do not include 0 are highlighted in bold. Abbreviations: CI – confidence interval.

**Supplementary Figure 1.** Completeness of the analyzed geolocation data for each hour in a day.

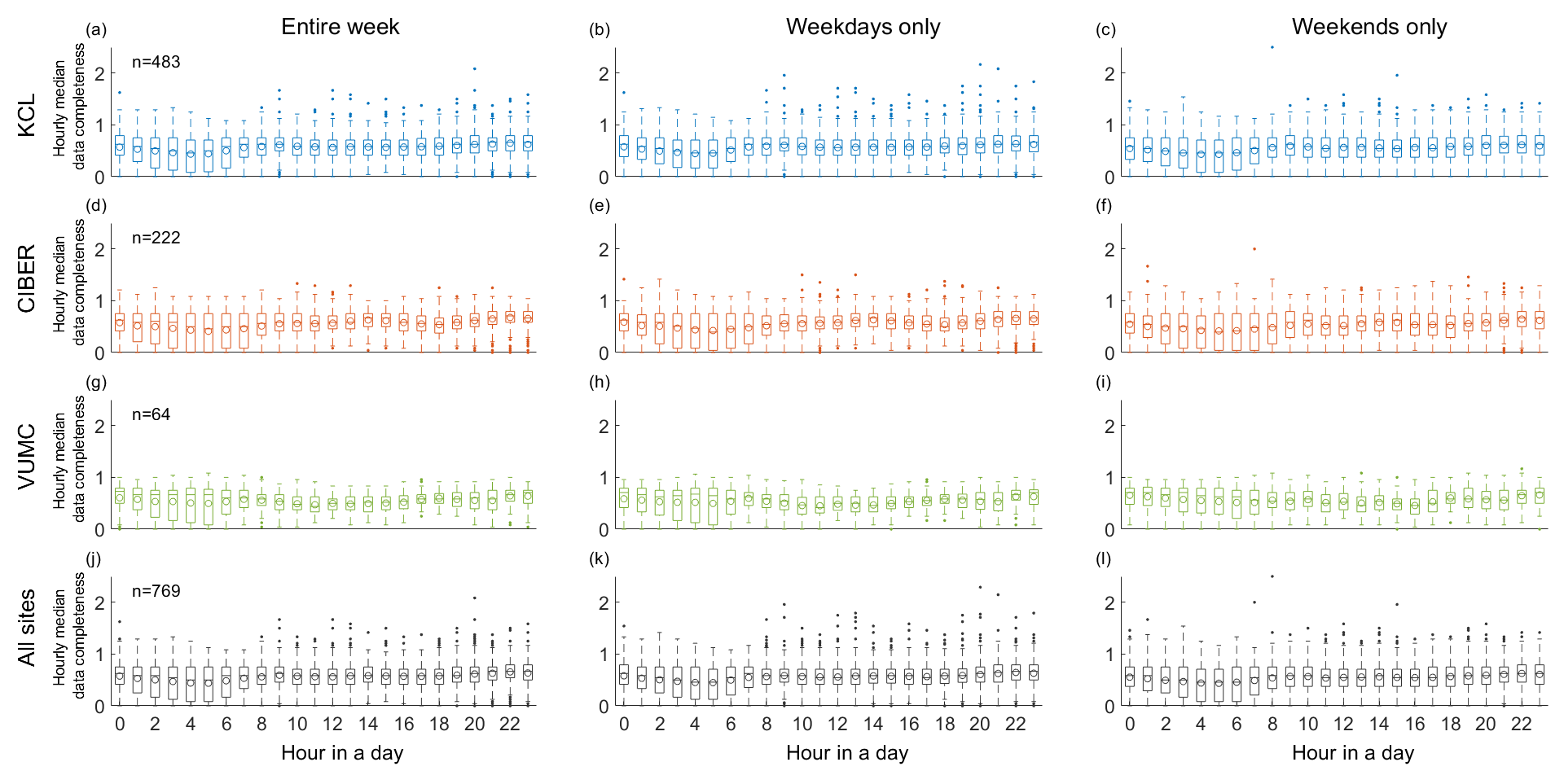

Data completeness is assessed for each clinical site (KCL: **a**, **b**, **c**; CIBER: **d**, **e**, **f**; VUMC: **g**, **h**, **i**; across the three sites: **j**, **k**, **l**) and analyzed time frame separately (the entire week: **a**, **d**, **g**, **j**; weekdays only: **b**, **e**, **h**, **k**; weekends only: **c**, **f**, **i**, **l**). Hour *H* along the x-axis corresponds to the geolocation data collected from *H*:00:00 to *H*:59:59. For each biweekly segment, data completeness for a given hour was computed as median daily completeness of the geolocation data collected during that hour across 14 days of the biweekly segment. *n* indicates the number of analyzed biweekly segments (Table 1). A horizontal bar and a circle in each boxplot indicate median and mean of the corresponding data, respectively. Data of different clinical sites are highlighted in different colors: KCL – light blue, CIBER – orange, VUMC – green, across the three sites (labeled as “All sites”) – black.

**Supplementary Figure 2.** Distributions of dataset characteristics for each clinical site separately.

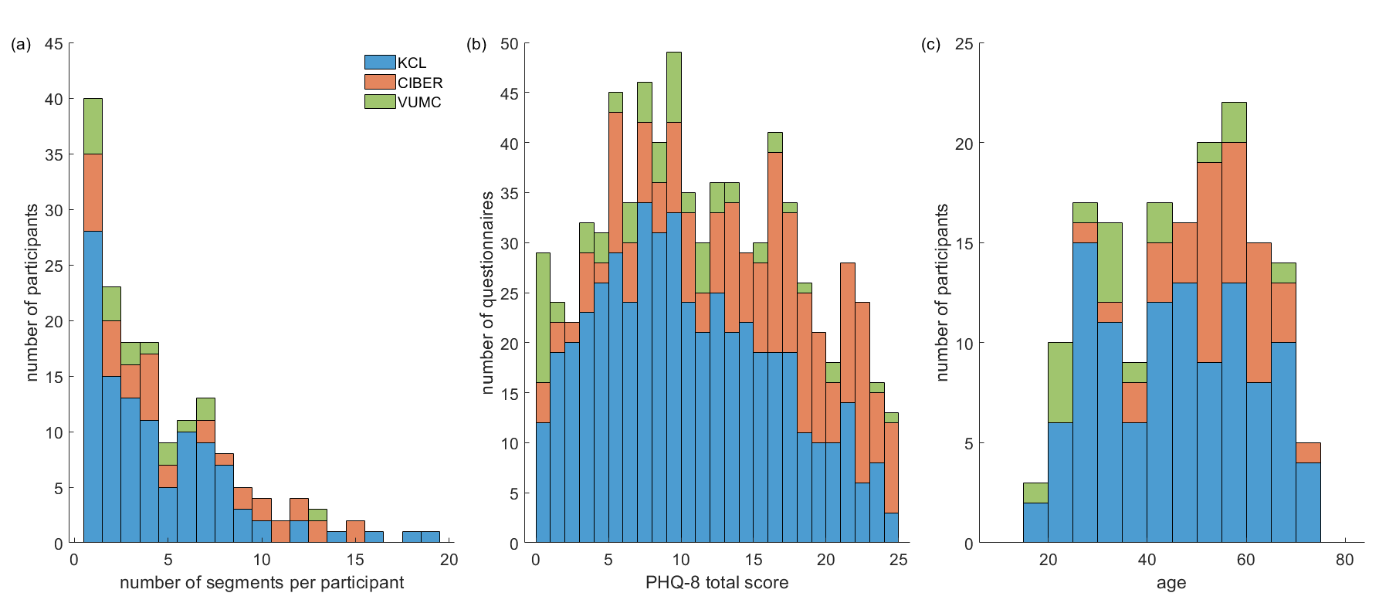

(**a**) Number of biweekly segments available for analysis per study participant. (**b**) PHQ-8 total score. (**c**) Participant age. The same data and abbreviations as in Figure 2 are used. Data of different clinical sites are highlighted in different colors: KCL – light blue, CIBER – orange, VUMC – green. Statistics on each presented dataset characteristic is reported in Table 1.

**Supplementary Figure 3.** Distributions of dataset characteristics for the employed versus unemployed participants.

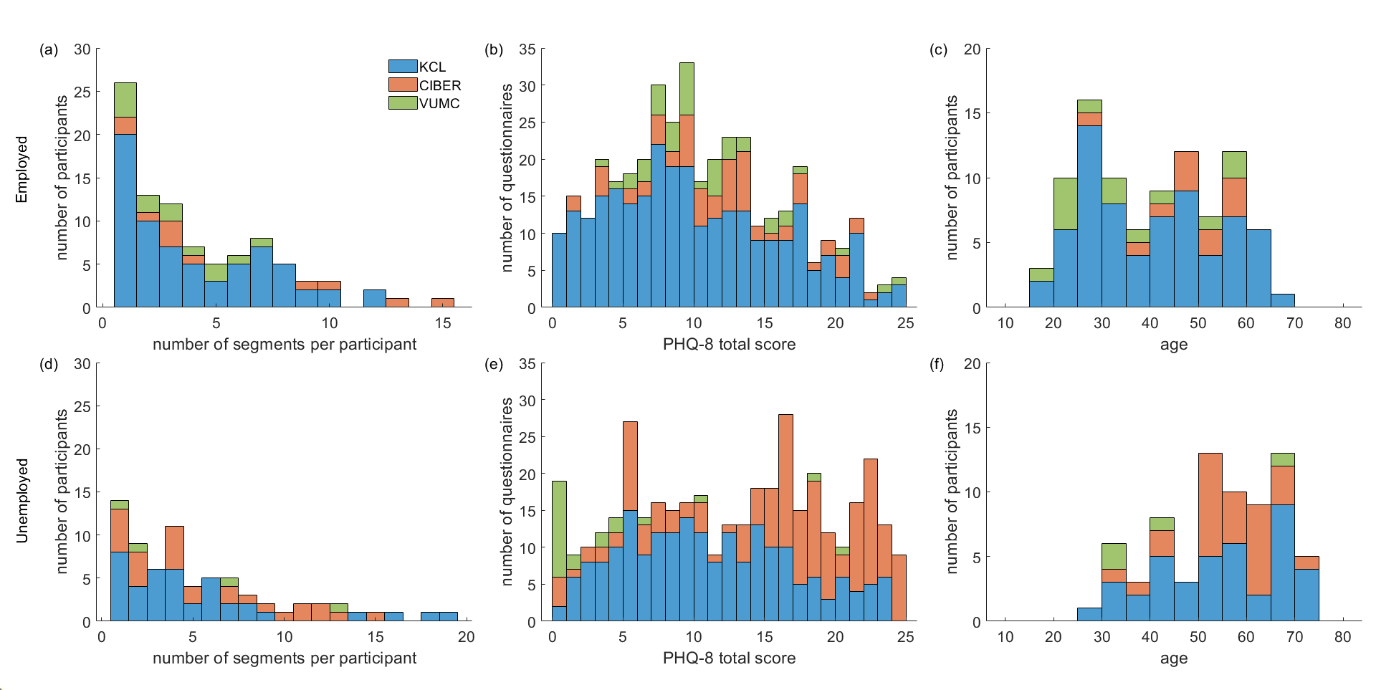

Data of the employed and unemployed participants are presented in the upper and lower rows, respectively. (**a, d**) Number of biweekly segments available for analysis per study participant. (**b, e**) PHQ-8 total score. (**c, f**) Participant age. Data of different clinical sites are highlighted in different colors: KCL – light blue, CIBER – orange, VUMC – green.

**Supplementary Figure 4.** Home stay computed (**a**) over the entire week, (**b**) for weekdays and (**c**) weekends only for each clinical site separately.

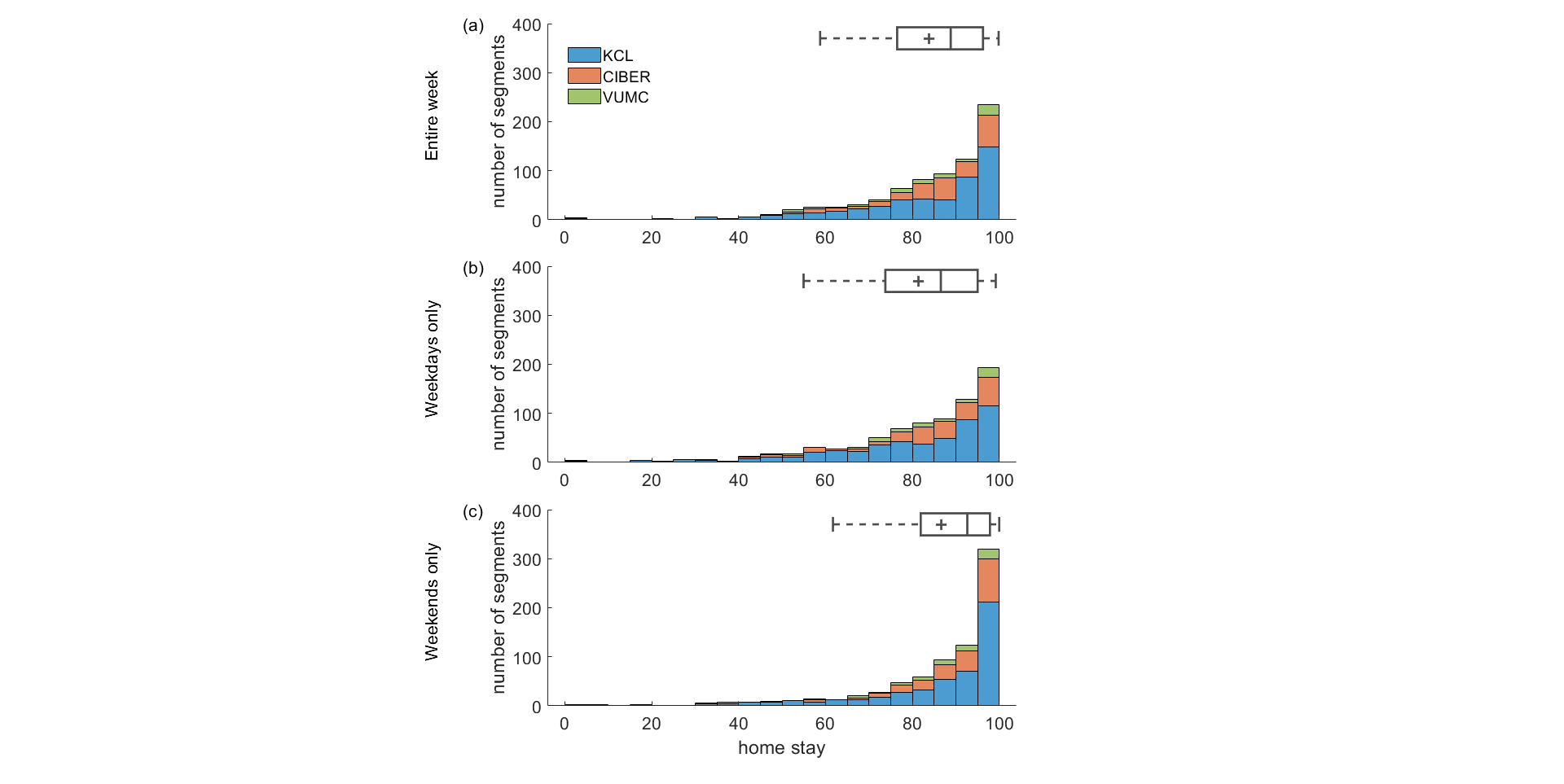

A grey horizontal bar and a cross in each boxplot indicate median and mean of the data pooled across the three sites, respectively. The same data and abbreviations as in Figure 3 are used. The data of different clinical sites are highlighted in different colors: KCL – light blue, CIBER – orange, VUMC – green.

**Supplementary Figure 5.** Distributions of home stay for the employed versus unemployed participants.

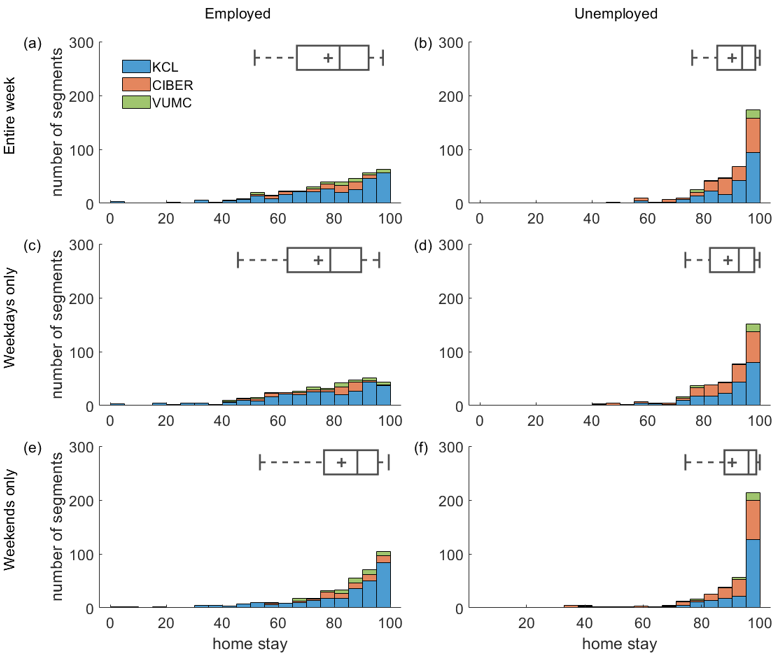

Data of the employed and unemployed participants are presented in the left and right columns, respectively. Home stay was computed (**a, b**) over the entire week, (**c, d**) for weekdays and (**e, f**) weekends only. A grey horizontal bar and a cross in each boxplot indicate median and mean of the data pooled across the three sites, respectively. The data of different clinical sites are highlighted in different colors: KCL – light blue, CIBER – orange, VUMC – green.
